# The Challenge of Forecasting Demand of Medical Resources and Supplies During a Pandemic: A Comparative Evaluation of Three Surge Calculators for COVID-19

**DOI:** 10.1101/2020.09.29.20204172

**Authors:** A.A. Kamar, N. Maalouf, E. Hitti, G. El Eid, H. Isma’eel, I. H. Elhajj

## Abstract

Ever since the World Health Organization (WHO) declared the new coronavirus disease 2019 (COVID-19) as a pandemic, there has been a public health debate concerning medical resources and supplies including hospital beds, intensive care units (ICU), ventilators, and Protective Personal Equipment (PPE). Forecasting COVID-19 dissemination has played a key role in informing healthcare professionals and governments on how to manage overburdened healthcare systems. However, forecasting during the pandemic remained challenging and sometimes highly controversial. Here, we highlight this challenge by performing a comparative evaluation for the estimations obtained from three COVID-19 surge calculators under different social distancing approaches, taking Lebanon as a case study. Despite discrepancies in estimations, the three surge calculators used herein agree that there will be a relative shortage in the capacity of medical resources and a significant surge in PPE demand as the social distancing policy is removed. Our results underscore the importance of implementing containment interventions including social distancing in alleviating the demand for medical care during the COVID-19 pandemic in the absence of any medication or vaccine. It is said that “All models are wrong, but some are useful,” in this paper we highlight that it is even more useful to employ several models.

## Introduction

The outbreak of the new coronavirus disease 2019 (COVID-19), caused by the severe acute respiratory syndrome coronavirus 2 (SARS-CoV-2), started in Wuhan City, China, and has rapidly spread all over the world. The newly identified coronavirus has 96% homology with the genetic sequence of β-coronaviruses; which include SARS-CoV and the Middle East respiratory syndrome CoV (MERS-CoV)^1,2^. Despite similarities, SARS-CoV infection was limited to specific geographic areas, while COVID-19 has been declared as a global pandemic by the WHO on 11 March 2020. As of 17 September 2020, there are 29,737,453 COVID-19 confirmed cases, 937,391deaths, and around 213 countries or territories affected with cases around the world^3^.

COVID-19 has not only caused mortality but has put tremendous pressure on healthcare systems and led to shortages in Personal Protective Equipment (PPE) ^4,5^. In the Middle East and North Africa Region (MENA) countries including Lebanon, the COVID-19 pandemic is leading to a health crisis and worsening the spike in demand on healthcare resources and supplies ^6^. To monitor the strain on healthcare systems in each country, it is essential to closely follow the demand for hospital beds, intensive care unit (ICU) beds, and ventilators ^7^. Therefore, a growing number of models have been established around the world to aid in forecasting COVID-19 deaths, cases, and the demand for medical supply (including ventilators, hospital beds, and ICU beds, the timing of patient surges, and more)^8–10^. While simple models apply the user’s inputs on the local population and current status of COVID-19, more sophisticated models permit the user to modify other parameters (e.g., social distancing changes) that affect the trends. Some of the models match projected cases to existing capacity to project when and where a caseload surge will surpass capacity^11^. Data-driven modeling approaches have also appeared for the COVID-19 pandemic, in which statistical and machine learning models are used for projecting cases, hospitalization, deaths, and the impact of social distancing^12^ Unfortunately, forecasting what is most likely to occur in the upcoming weeks during the COVID-19 pandemic is not available for all countries including some European countries or few states in the USA ^8^. Where available, different models are providing widely varying numbers of needed medical resources and/or supplies which often lead to an incorrect distribution of what is available due to inconsistency in numbers. For example, the Centers for Disease Control and Prevention has reported that COVID-19 outbreaks in parts of the USA have resulted in surges in hospitalizations and ICU patients ^13^. However, providing accurate predictions of the healthcare system capacity peak demand is controversial due to the scarcity and/or unreliability of data in addition to challenges associated with forecasting the effects of the rapid changes in mitigation policies ^14^. So far, the efforts to accurately model any emerging outbreak’s trajectory for the upcoming days are limited due to variabilities in assumptions and parameters including social distancing^14–16^. Accordingly, the use of a single forecasting model may not precisely predict how the pandemic evolves ^17^.

Regardless of all the challenges, COVID-19 has put forecasting at the top of global public policymaking and developing effective preventive strategies ^18,19^. Since there is no “gold standard” for predicting thresholds, the reasonable evaluation of the outputs of various forecasting models has remained an open question ^20^. In this work, we highlight this challenge by comparing the projected demand for medical resources and supplies from three surge calculators for COVID-19 taking Lebanon as our case study. To this end, we adapted an available statistical model for estimating the daily impact of COVID-19 on hospital services based on the COVID-19 Hospital Impact Model for Epidemics (CHIME). The model was modified to incorporate longer projection periods and different social distancing policies (%). We then compared the hospital beds, ICU beds, and PPE demand over 200 days projected by the three surge calculators a) CHIME PPE calculator, b) WHO COVID-19 Essential Supplies Forecasting Tool (COVID-19 ESFT) and c) our own developed American University of Beirut Medical Center (COVID-19 AUBMC Surge needs) calculator. The results of these calculators differ depending on the parameters and assumptions implemented within to generate the forecasted data. Despite the discrepancies in estimations, the three surge calculators used herein consistently agree on trends demonstrating that social distancing policy can help reduce the demand for medical resources and supplies amid the COVID-19 pandemic.

## Methods

### Data collection

#### Case counts, population and hospital capacity data

We used the WHO and the Lebanese Ministry of Public Health (MoPH) websites to identify data on the estimated Lebanese population in 2020 (6,825,000) and confirmed COVID-19 infected individuals and death cases respectively. Data on inpatient beds, ICU capacity, and mechanical ventilators were obtained from the Lebanese MoPH website (Table 1). The data for the COVID-19 AUBMC calculator regarding PPE demand were collected based on historical and current data estimates from the American University of Beirut Medical Center (AUBMC), presuming these data as a nationwide reference. In most of the calculations, numbers derived from Lebanon were used where available.

**Table 1.** Input parameters used for forecasting healthcare demand in Lebanon during the COVID-19 pandemic through CHIME Model/calculator, COVID-19 AUBMC, and WHO COVID-19 ESFT calculators.

| Parameters | Value | Reference/Source |
| --- | --- | --- |
| Lebanese population | 6,825,000 | WHO COVID-19 ESFT Calculator <sup>28</sup> |
| Currently hospitalized COVID-19 patients | 67 | MOPH report 14/5/2020 |
| Doubling time before social distancing (days) | 4 days | calculated from the days following the first coronavirus case in Lebanon on February 21, 2020, https://www.moph.gov.lb/maps/covid19.php |
| Hospitalization Percentage in Lebanon (%) | 20 % | https://www.moph.gov.lb/en/Pages/2/24870/novel-coronavirus-2019- |
| Average Days in ICU | 9 | MoPH report 27/3/2020 |
| ICU Percentage (%) | 5 % | https://www.moph.gov.lb/en/Pages/2/24870/novel-coronavirus-2019- |
| Mechanical Ventilators Percentage (%) | 3 % | https://www.moph.gov.lb/en/Pages/2/24870/novel-coronavirus-2019- |
| Hospital Length of Stay (days) | 28 | http://gabgoh.github.io/COVID/index.html |
| Hospital Market share (%) | 100 % | The percentage of patients in the region that are expected to come to a specific hospital (as opposed to other hospitals in the region) when they get sick<sup>21</sup>.. |
| Infectious days | 14 | CHIME Model <sup>24</sup> |
| Number of Projected Days | 200 | - |
| Currently Infected individuals (as of March 27<sup>th</sup>, 2020) | 391 | MOPH report 27/3/2020 |

## Statistical Input parameters

### CHIME Model and modifications

Our estimations were based on the CHIME model that was initially developed by the Predictive Health at Penn Medicine in the USA^21^, which permits healthcare systems to enter data about their population and modify the assumptions about the COVID-19 spread and behavior^21^. The tool runs a modified Susceptible-Infected-Recovered (SIR) model, a traditional epidemiological forecasting technique, to estimate the number of new COVID-19 hospital admissions per day. The SIR model calculates the theoretical number of people infected with an infectious disease over a period of time in a closed population with the Kermack-McKendrick model being the simplest of all SIR models^22,23^. The CHIME model was built to help hospital systems to accurately project the needed resources during the COVID-19 pandemic, mainly hospital beds, ICU beds, and ventilators ^24^. The model accounts for four main parameter categories: 1) the hospital parameters, 2) spread and contact parameter, 3) severity parameters, and 4) display parameters. The model allows integrating some information about social distancing policies that are emplaced by governments ^24^.

In this study, we have introduced some modifications to the CHIME application to make it compatible with the input parameters available for the projections in Lebanon.

The modifications were twofold:

#### 1. Automate the data projection process

The application was updated allowing the user to enter the input parameters into an “Excel workbook” and obtain the generated census compiled together in the same workbook. This alleviates the data processing part where the user needs to obtain each projection separately. It also enables faster simulations for different input parameters and most importantly different social distancing measures (Table 2).

**Table 2.**
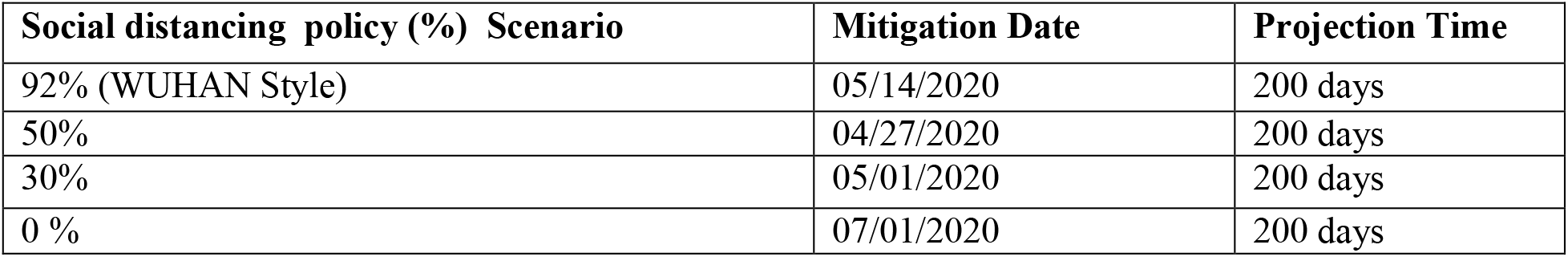
The four different social distancing policy scenarios presumed, along with their subsequent mitigation dates, to estimate the capacity of hospital beds, ICU beds, ventilators, and PPE demands during the COVID-19 pandemic in Lebanon over 200 days.

### 2. Allow projections over a longer period

The application initially allowed for a maximum 30-day projection period. Our contribution enables for longer periods such as 200 days, as seen herein. This provides analysts and healthcare workers with longer forecasts, thus giving them more time to prepare for periods of peak demand. The CHIME projects the daily and cumulative number hospitalized, ICU, ventilated, and newly admitted COVID-19 cases, with social distancing policy percentage being the variable.

#### 3. Provide simultaneous known cases (per day) as output

The modification allows us to study the number of daily infected individuals after deducting the number of recovered cases. The number reflects the confirmed cases and does not consider possible infected individuals that are not confirmed yet. The aim behind this modification is to provide these numbers as inputs to the calculators, especially COVID-19 AUBMC and WHO COVID-19 ESFT calculators.

Our projections were based on three social distancing policy scenarios (0%, 30%, and 50%) based on the National Health Strategic Preparedness and Response Plan for COVID-19 pandemic lock-down management and exit strategy implemented by the Lebanese Government ^6^. We also simulated the strict social distancing policy (92%) based on the Wuhan-style containment ^25^. The latter was performed to assess how strict “lock-down” could help better contain the COVID-19 outbreak and maintain the demand for hospital beds, ICU beds, ventilators, and PPE. Although Lebanon started re-opening gradually with limited capacity as of May 4, 2020, the country went again into a complete four-day lockdown starting May 14. Therefore, we ran our estimations on May 14, 2020 (Day 0) assuming the four social distancing policy scenarios. The other social distancing policy scenarios and the mitigation dates below were chosen as the per five-step re-opening plan implemented by the Lebanese Government ^6^(Table 2).

### PPE calculators

We have used the WHO COVID-19 ESFT calculator (WHO 2020a) and the COVID-19 AUBMC surge Needs calculator, to estimate daily and total hospital beds, ICU beds, ventilators, and PPE demand. Results obtained from the CHIME PPE calculator were then compared to those derived from the WHO COVID-19 ESFT and COVID-19 AUBMC calculators. As the COVID-19 AUBMC calculator was developed by our team, we chose the CHIME PPE calculator because it is compatible with the CHIME model, and the WHO COVID-19 ESFT calculator because it is widely used as a worldwide reference to estimate medical resources and supplies to respond to the current COVID-19 pandemic. Also, the CHIME and COVID-19 ESFT calculators are freely available online for use by governments, stakeholders, and healthcare centers.

#### The CHIME PPE calculator

The CHIME PPE calculator was generated to work in parallel with CHIME-generated projections. The calculator uses forecasted patient censuses to output daily and cumulative projections for each type of PPE (including N95 masks, surgical masks, gloves, gown, and eye disposable protection) quantities per day, and computes the cumulative PPE predictions ^26^. This tool also permits users to input their custom scenarios (standard, crisis, contingency, and custom), tailored to the specific situations relevant to their hospital or healthcare system ^26^. In our study, we chose the values for the standard scenario assuming we do not have exact publicly available data estimates of the exact number of staff and HCW (healthcare workers) in Lebanon.

As social distancing policies and gradual lifting of restrictions are always the keep factors being debated in the intervention strategies, our estimations were based on the four social distancing policy scenarios mentioned above (Table 2). For each scenario, the projected hospitalized, ICU, and ventilated censuses are inserted into the CHIME PPE calculator in addition to the daily admissions. This data is obtained from the Penn CHIME application model. Based on the provided data the calculator outputs the PPE predictions per day in addition to the total cumulative predictions over the chosen period of 200 days. Since other calculators do not forecast the daily PPE demand, we compared the average daily PPE demand from this calculator with the predictions of other calculators.

#### WHO COVID-19 ESTF Calculator

The 2020 WHO COVID-19 ESTF calculator is developed to aid countries in estimating potential needs and supplies to respond to the COVID-19 pandemic ^27^. The tool is not meant to be used as an epidemiological model, yet it has simple exponential growth and Susceptible-Infectious-Removed (SIR) case forecast options built-in ^28^. The calculator is a supply forecasting tool that helps in estimating potential requirements for essential supplies including PPE (e.g. surgical masks, gloves, gowns, goggles, respirators, and face shields), biomedical equipment for case management (e.g. mechanical ventilators and oxygen concentrators), drugs for supportive treatment, hygiene, and IPC commodities, diagnostics, and consumable medical supplies. The calculator also estimates the weekly number of COVID-19 patients classified according to severity as follows: mild, moderate, severe, and critical. Severe and critical cases are admitted to ICU and require oxygen and ventilation respectively. Inpatient beds in this calculator refer to ICU beds occupied by the critical and severe COVID-19 cases per week and not day. Therefore, the estimated peak for inpatient and ICU beds will be the same throughout the chosen period. Although the tool is suited for projections over a short period (12 weeks), it offers an option to enter data manually and make projections over longer periods. COVID-19-ESFT does not quantify or account for resources already available locally or those pending delivery. The calculator projects the PPE quantity per person per day for inpatient care, cases in isolation, screening, and laboratory, and the total daily costs (USD) of items over 28 weeks. Then, it adds the total quantity for each per day. In this study, we use the default input parameters set by the WHO COVID-19 ESFT calculator for Lebanon, including the population estimate, patients case sensitivity, healthcare workers, and staff. To maintain consistency across the different calculators, we manually input the cumulative projected COVID-19 cases as obtained from the CHIME model application. The data is compiled in a weekly form (up to 28 weeks) to be compatible with the COVID-19 ESTF calculator. This approach guarantees that all calculators are using the same numbers for infections and the focus would be on the discrepancies/agreements between the models on the estimates of resource demand.

#### The COVID-19 AUBMC surge needs calculator

The AUBMC calculator (https://www.aub.edu.lb/fm/vmp/Pages/calculators.aspx) was developed by our team based on AUBMC and MoPH data and is implemented as an excel file that predicts the average total of PPE needs (e.g., gloves, surgical face masks, face shields, and N95 masks) per day. The estimations are based on the average number of patients admitted and tested individuals per day using actual PPE demand data collected from AUBMC. Note that we assume AUBMC policies of PPE use (such as wearing surgical masks over N95 masks, replacing surgical masks per patient, and replacing N95 masks per shift) are representative of policies at the national level. The average number of admitted patients is calculated from the Penn CHIME model application projections over the whole pandemic period (200 days) using the hospitalization percentage of simultaneously infected individuals. The average number of tested individuals is calculated by dividing the total number of tests done as indicated in (https://www.worldometers.info/coronavirus/?#countries) by the number of pandemic days. The PPE estimations in this calculator are based on average values as compared to daily/weekly projections used in the other two calculators. This calculator also projects the total number of ventilators, hospital beds, and ICU beds occupied during the COVID-19 pandemic using the same inputs of the CHIME application. The beds and ventilators calculations are designed to estimate the availability during peak periods of the pandemic. The calculator takes as input the currently available ventilators and beds dedicated to COVID-19 patients as declared by MoPH in addition to the peak simultaneous infections and admissions as projected by the Penn CHIME model application. This information helps in forecasting possible shortages in beds or ventilators and the dates when these shortages would occur (Table 3).

**Table 3.**
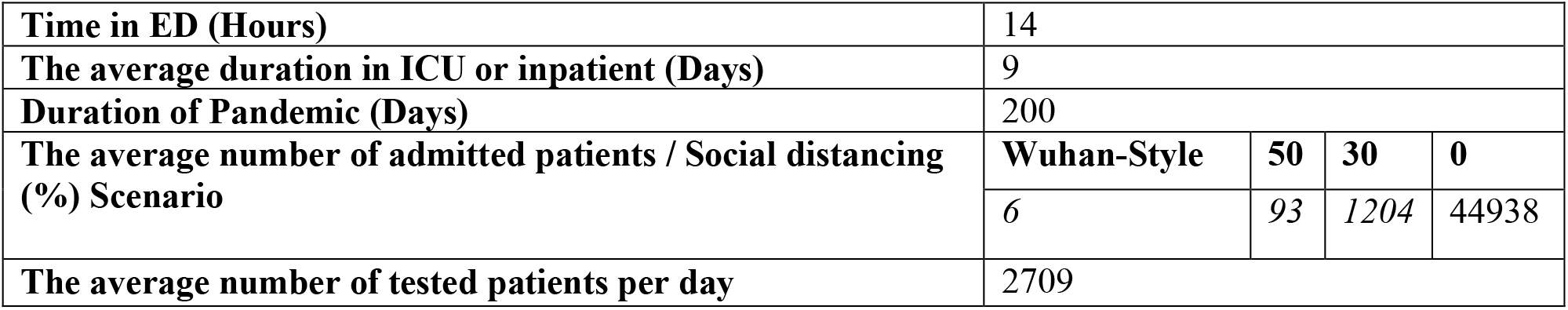
Data assumptions used to estimate peak daily hospital beds, ventilators, and the average daily PPE demand using the COVID-19 AUBMC surge needs calculator.

|  |  |  |  |  |
| --- | --- | --- | --- | --- |
| Time in ED (Hours) | 14 |  |  |  |
| The average duration in ICU or inpatient (Days) | 9 |  |  |  |
| Duration of Pandemic (Days) | 200 |  |  |  |
| The average number of admitted patients / Social distancing (%) Scenario | Wuhan-Style | 50 | 30 | 0 |
|  | 6 | 93 | 1204 | 44938 |
| The average number of tested patients per day | 2709 |  |  |  |

## Results

Based on MoPH data, from March 29, 2020, Lebanon is estimated to have 2308 ICU beds and 11794 inpatient (hospital) beds. We assume that 32 % of ICU beds and 20% of hospital beds at the national level are currently occupied by non-COVID-19 patients (Table 4). However, this occupancy number is, in reality, higher due to the casualties from the devastating port explosion in Beirut on August 4, 2020. Also, there has been a surge in cases nationally. One reason behind this surge is that many people have been unable to follow precautionary measures, such as social distancing, during the relief efforts in Beirut. Also, some of the major hospitals have been partially or heavily damaged as a result of the huge blast at Beirut’s port. MoPH reported that almost all COVID-19 beds are full and hospitals are running out of space for new patients by the time of writing this work. This incident highlights the challenge of planning during a pandemic, particularly when coupled with unexpected disasters.

**Table 4.** The estimated total and available numbers of hospital beds, ICU beds, and ventilators for the COVID-19 and non-COVID-19 patients in Lebanon according to the MoPH daily report on March 29, 2020

| Hospital Beds | Total Number | % of beds in use (non-COVID-19 Patients) |  | Number Available (COVID-19 patients) |
| --- | --- | --- | --- | --- |
| In-patient | 15195 | 20 |  | 9435 |
| ICU | 2308 | 32 |  | 1569 |
| Ventilators (Infant and adult) | 1185 | % Out of Service | % in Use (non-COVID-19) | 687 |
|  |  | 2 | 40 |  |

In one scenario and based on our data, assuming strict social distancing policy or the Wuhan-style (92%), the modified CHIME model estimates that out of the remaining 9435 hospital beds, a peak of 67 inpatient beds is needed by the COVID-19 cases or 0.7 % of the available hospital beds. Of the remaining available 1569 ICU beds and 687 ventilators, the model estimates a peak of 15 (1%) ICU beds, and 10 ventilators (1.5%) are needed by COVID-19 patients respectively, with a decreasing pattern in the estimated COVID-19 infected patients over the projected period. Assuming the same scenario, the COVID-19 ESFT calculator estimates a weekly peak of 11 inpatient beds (0.1%), 17 ICU beds (1.1%), and 9 ventilators (1.3%) are occupied. The COVID-19 AUBMC calculator estimates a different peak of 71 inpatient beds (0.8%), 18 ICU beds (1.1%), and 11 ventilators (1.6 %) (Figs. 1.A, 2, and 3). Upon relaxing social distancing measures to 50 %, the CHIME model estimates the same peak capacity for the available inpatient beds, ICU beds, and ventilators. Still, the COVID-19 AUBMC calculator differentially estimates that the peak of daily occupied inpatient beds increases to 94 (1%), while ICU beds and ventilators increase to 23, representing 1.47% and 3.35% of the available ICU beds and ventilators respectively. On the other hand, the COVID-19 ESFT estimates an increase in the weekly peak demand of inpatient bed capacity to 1279 (13.75%), the ICU beds to 58 (3.7%), and the ventilators to 16 (1.31%) (Figs. 1.B, 2, and 3).

**Figure 1.**
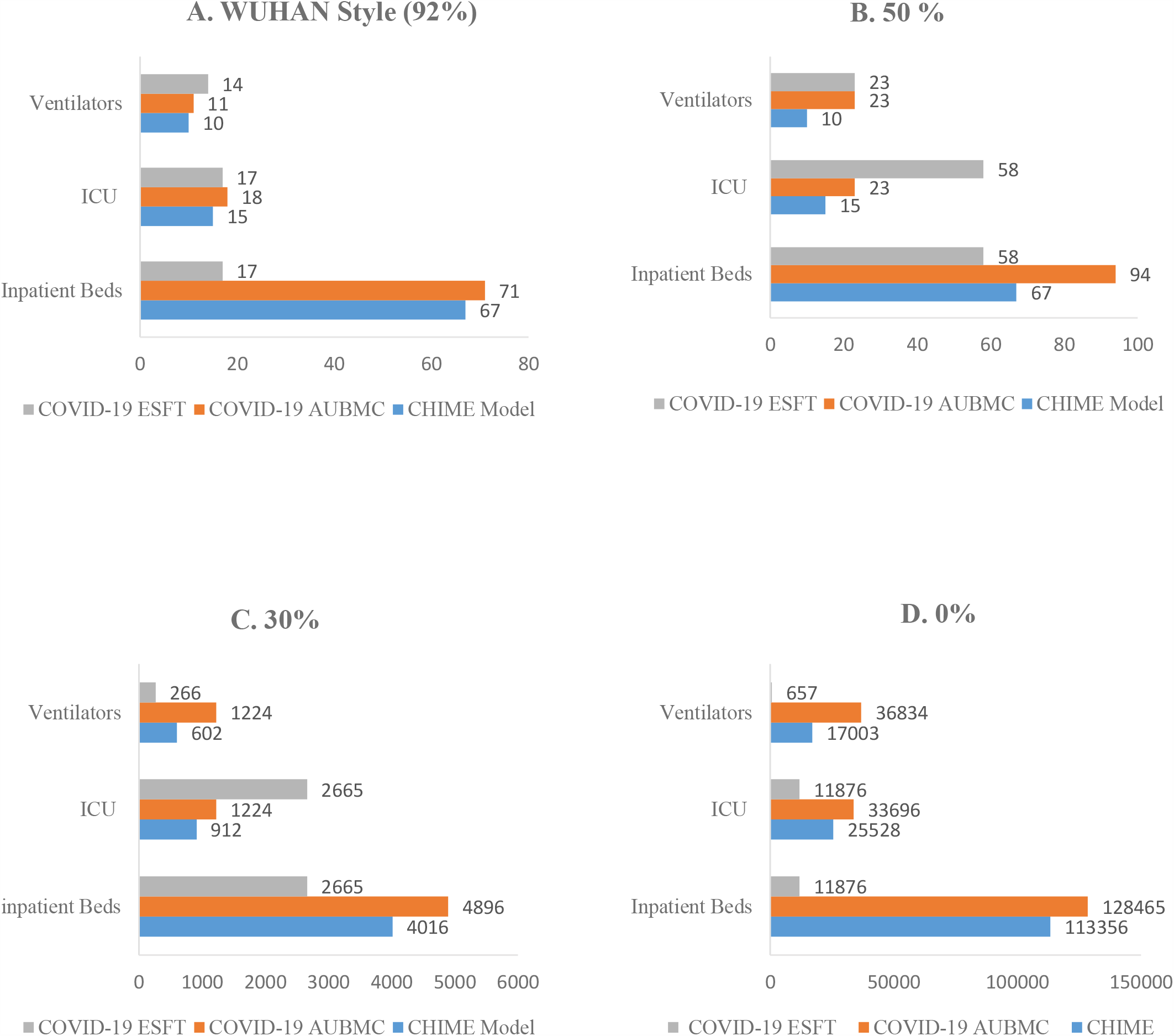
The forecasted peak of the daily inpatient beds, ICU beds, and ventilators by the three surge calculator during the COVID-19 pandemic in Lebanon over 200 days assuming the four social distancing policy scenarios (A, B, C, and D).

When the social distancing policy is reduced to 30 %, all the surge calculators variably show that the capacity of the daily peak of hospital beds and ventilators occupied by COVID-19 patients significantly increases. The CHIME estimates a daily peak of 4016 inpatient beds (43%), 912 ICU beds (58%), and 602 ventilators (88%) to be occupied. With a higher occupancy rate, the COVID-19 AUBMC calculator estimates a peak of 4896 inpatient beds (52%), 1224 ICU beds (78%), and 1224 ventilators. The latter indicates a shortage of 537 ventilators for COVID-19 patients in this scenario. On the contrary, the ESFT calculator estimates a marked increase in the weekly peak of inpatient beds (2665) and ventilators (266) but a shortage of 1096 ICU beds (Figs. 1.C, 2, and 3). To evaluate to which extent removing social distancing practices strain hospital capacity, we ran our estimations assuming 0 % social distancing. Our results show that in addition to the variability in projections, all calculators estimated that there will be a shortage in inpatient and ICU bed capacity during the 200 days. Yet, only ESFT calculator estimates that ventilators available can accommodate the critical COVID-19 patients needing ventilation. The CHIME model estimates that the daily peak capacity significantly increases to 113356 inpatient beds, 25528 ICU beds, and 17003 ventilators. Similarly, this increase was marked by the COVID-19 AUBMC calculator that estimated a peak daily capacity of 124658 inpatient beds, 33696 ICU beds, and 36834 ventilators. The ESFT calculator forecasted a daily peak capacity of 11876 inpatient beds, 11876 ICU beds, and 657 ventilators (Figs. 1.D, 2, and 3).

We also compared the projected demand for different PPE types (including N95, surgical masks, gloves, gowns, disposable eye protection/goggles, and face shield) by the three surge calculators over 200 days assuming the same social distancing policy scenarios (%). The CHIME calculator estimates a minimum average daily demand of various PPE types when Wuhan style (92%) policy is applied. The projected outputs include an average daily demand of 689 N95 masks, 1239 surgical masks, and 13086 gloves, gowns, and disposable eye protection. When compared to the CHIME projections, the COVID-19 AUBMC calculator estimates a slightly more daily average demand of 817 N95 masks, however, it estimates an average daily peak demand of 11516 surgical masks, 57842 gloves, and 11811 gowns, and 11429 face shields. Interestingly, the ESFT calculator estimates a significantly higher average daily demand values for all PPE types. The estimated average daily demand includes 75082 surgical masks, 159488 gloves, 42331 gowns, 36941 goggles, and160 face shields (Fig.4A). The estimated average daily demand for gloves in the ESFT calculator refers to the sum of gloves used for surgery, examination, and heavy-duty. When social distancing policy changes to 50%, the forecasted average daily demand for all PPE types increases gradually to 12023 N95 masks, 216411 surgical masks, 228434 gloves, gowns, and disposable eye protection. This increase in the projected average daily PPE demand was also observed with the COVID-19 AUBMC and ESFT calculators. The COVID-19 AUBMC estimates 883 N95 masks, 13201 surgical masks, 72317 gloves, 14274 gowns, and 11796 face shields. The ESFT estimates an average daily demand of 269432 surgical masks, 926610 gloves, 78503 gowns, 71158 goggles, and 280 face shields (Fig. 4B). The projected average daily demand for all PPE types by the three surge calculators was still revealing a spike in values with the 30 % social distancing policy. The CHIME calculated a total average daily demand of 84761 N95 masks, 1525695 surgical masks, 1610456 gloves, 1610456 gowns, and 1610456 disposable eye protection. AUBMC estimates 1716 N95 masks, 34705 surgical masks, 257035 gloves, 45709 gowns, and 16472 face shields. A significant surge in the average daily demand was also obtained with the COVID-19 ESFT calculator with an estimation of 2786405 surgical masks, 210525007 gloves, 535175 gowns, 510030 goggles, and 8201 face shields (Fig. 4C). On the other hand, the variability in projections was more emphasized when we applied the 0% social distancing policy. The CHIME projected estimated an average daily demand of 2701804 N95, 48632478 surgical masks, and 51334282 for gloves, gowns, and disposable eye protection. AUBMC estimated an average daily demand of 34517 N95 masks, 881405 surgical masks, 7530079 gloves, 1283394 gowns, and 200594 face shields. This trend was also obtained with COVID-19 ESFT, estimating an average daily need of 26385273 surgical masks, 45233595 gloves, 1982045gowns, 1248759 goggles, and 15433 face shields (Fig.4D).

**Figure 2.**
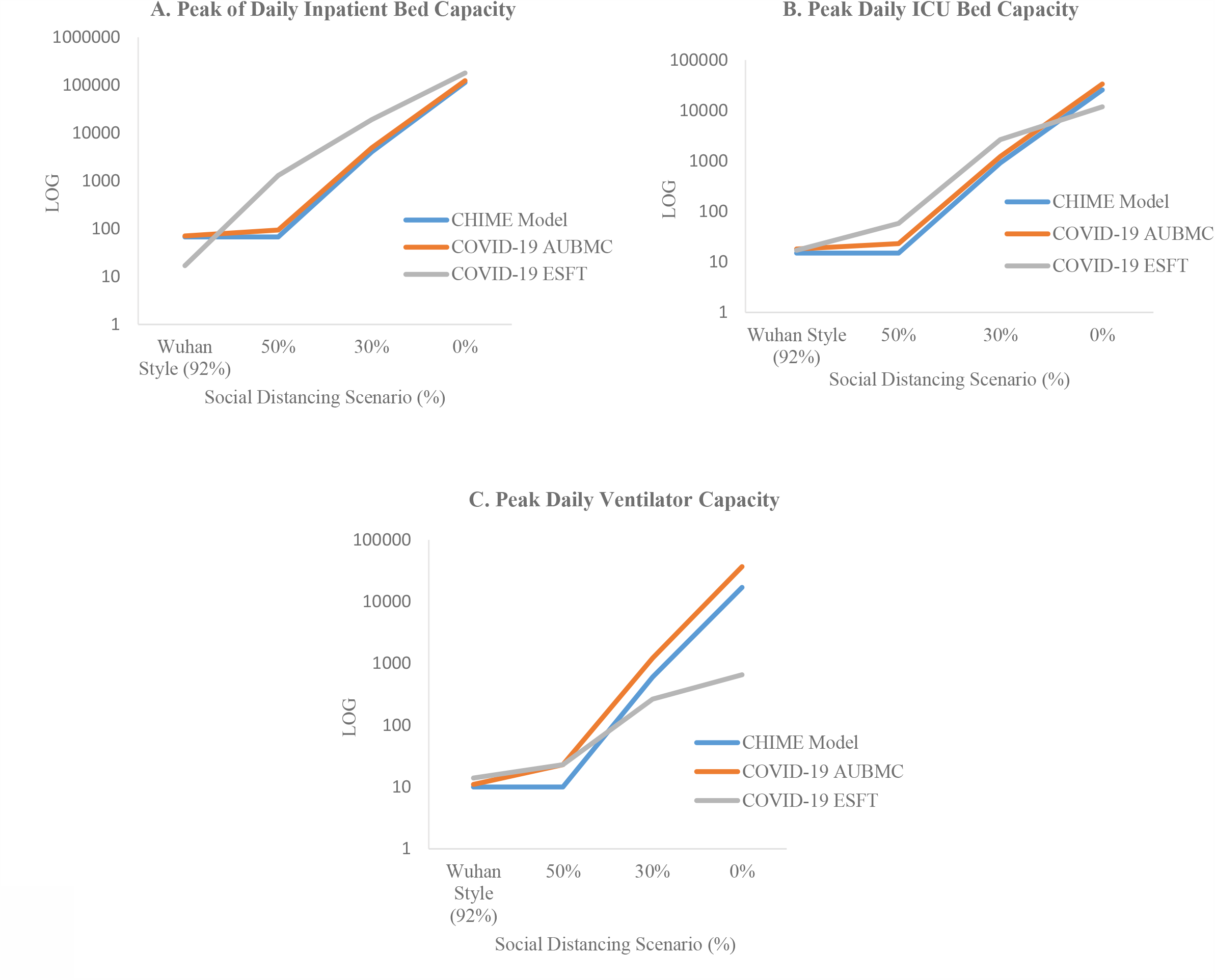
The variation in the forecasted peak capacity for inpatient beds (A), ICU beds (B), and ventilators (C) assuming the four social distancing policy using the three surge calculators over 200 days in Lebanon during the COVID-19 pandemic. All calculators show a significant increase in the number of occupied beds and ventilators upon relaxing social distancing measures.

**Figure 3.**
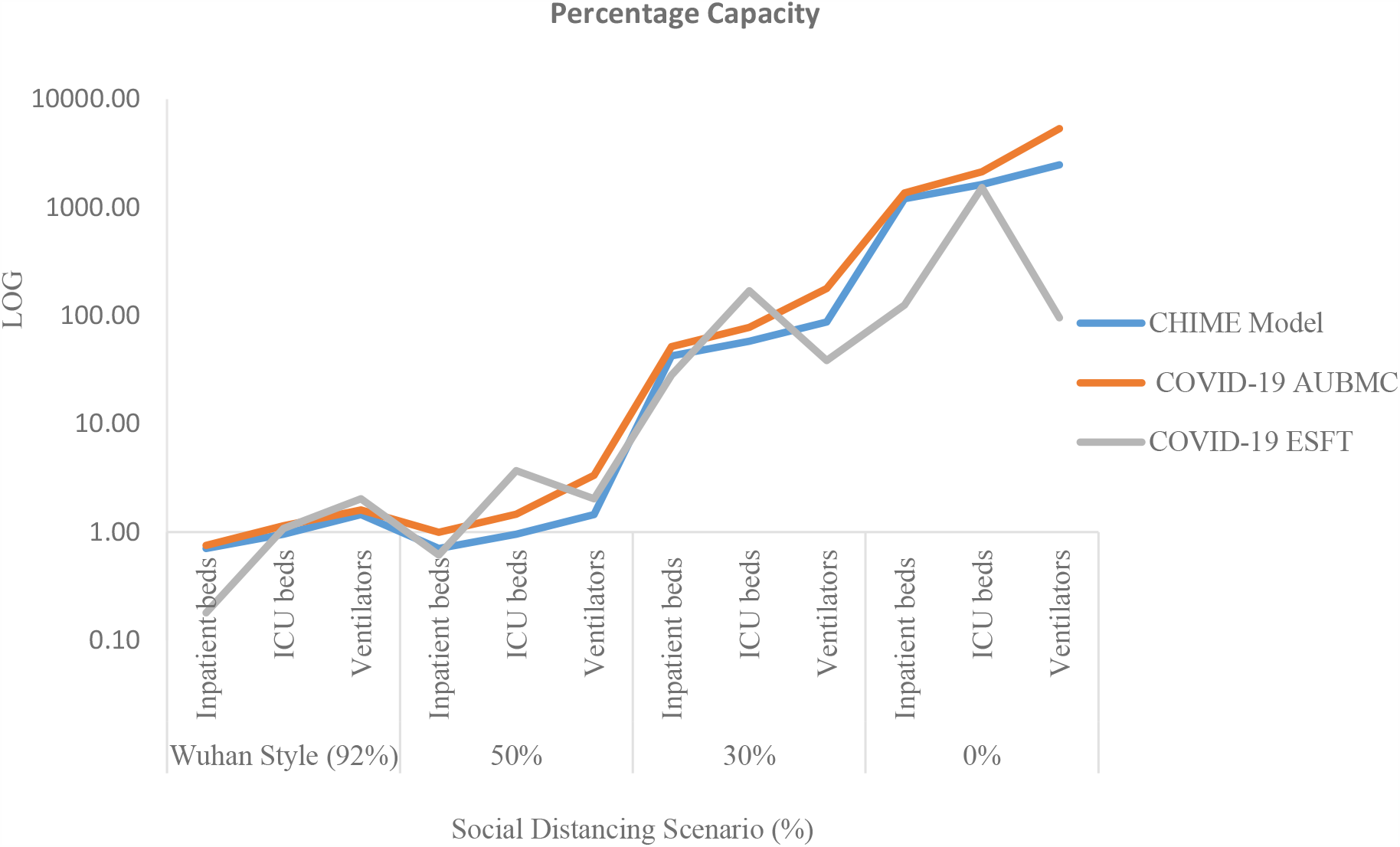
The variation in the projected percentage capacity of occupied hospital beds and ventilators upon changing the social distancing policy scenarios (%), using the three surge calculators during the COVID-19 pandemic in Lebanon. The three calculators forecast a sharp increase in the percentage of the occupied inpatient beds, ICU beds, and ventilators by the COVID-19 patients when social distancing measures are relaxed to 0%.

**Figure 4.**
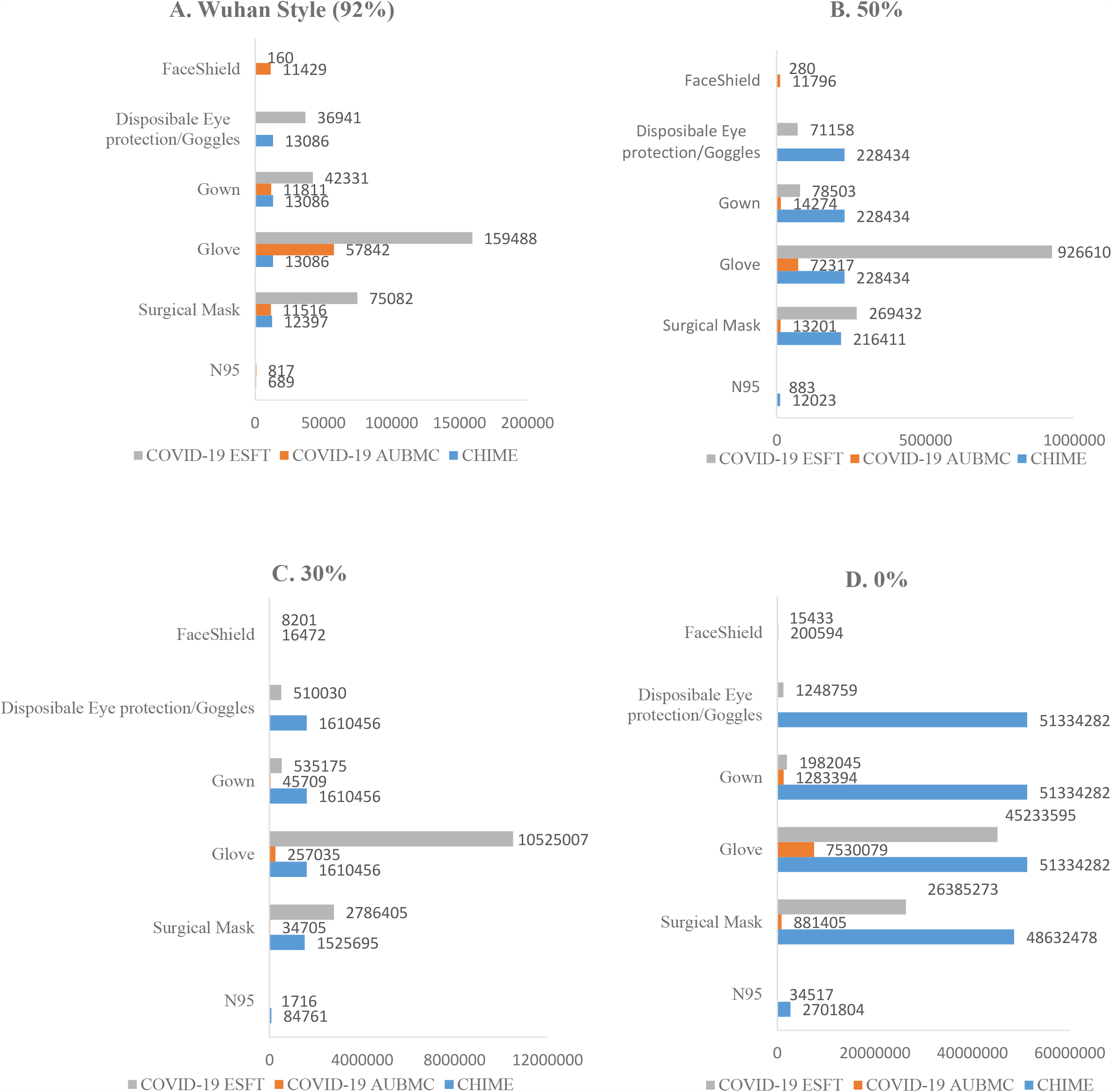
Comparing the forecasted average daily demand for all PPE types by the three surge calculators during the COVID-19 pandemic in Lebanon for 200 days, assuming the four social distancing policy scenarios (A, B, C, and D). The surge calculators forecast a significant spike in the average daily PPE demand as social distancing measures are relaxed.

We then compared the change in the forecasted PPE fold-demand from other scenarios with the Wuhan Style scenario (92%) since we have the least contact rate, minimum COVID-19 cases, and minimal projected hospital utilization and PPE demands. So, when we changed social policy to 50%, the CHIME estimated an average of a 17-fold increase in the daily demand for all the PPE types. However, the COVID-19 AUBMC and ESFT reveal a minimum of a 1- and 4-fold increase in the average PPE demand per day. This difference in PPE demand projections becomes more apparent as social distancing policy decreases to 30% where the ESFT and the CHIME estimate up to 46 and 100-fold increase in the average daily demand for all PPE types respectively. Although the COVID-19 AUBMC calculator projected a substantial increase in the average daily demand for all PPE types, this pattern in fold increase was not observed. The calculator, however, estimates only a 4-fold increase in the average daily demand for all PPE types. The difference in estimations was more obvious with the relaxed social distancing scenario (0%). While the CHIME estimates a significant increase in the average daily demand to 3000 folds for all PPE types, the COVID-19 AUBMC and ESFT forecast of around 100- and 200-fold increase in the average daily demand for all PPE types respectively (Fig. 5).

**Figure 5.**
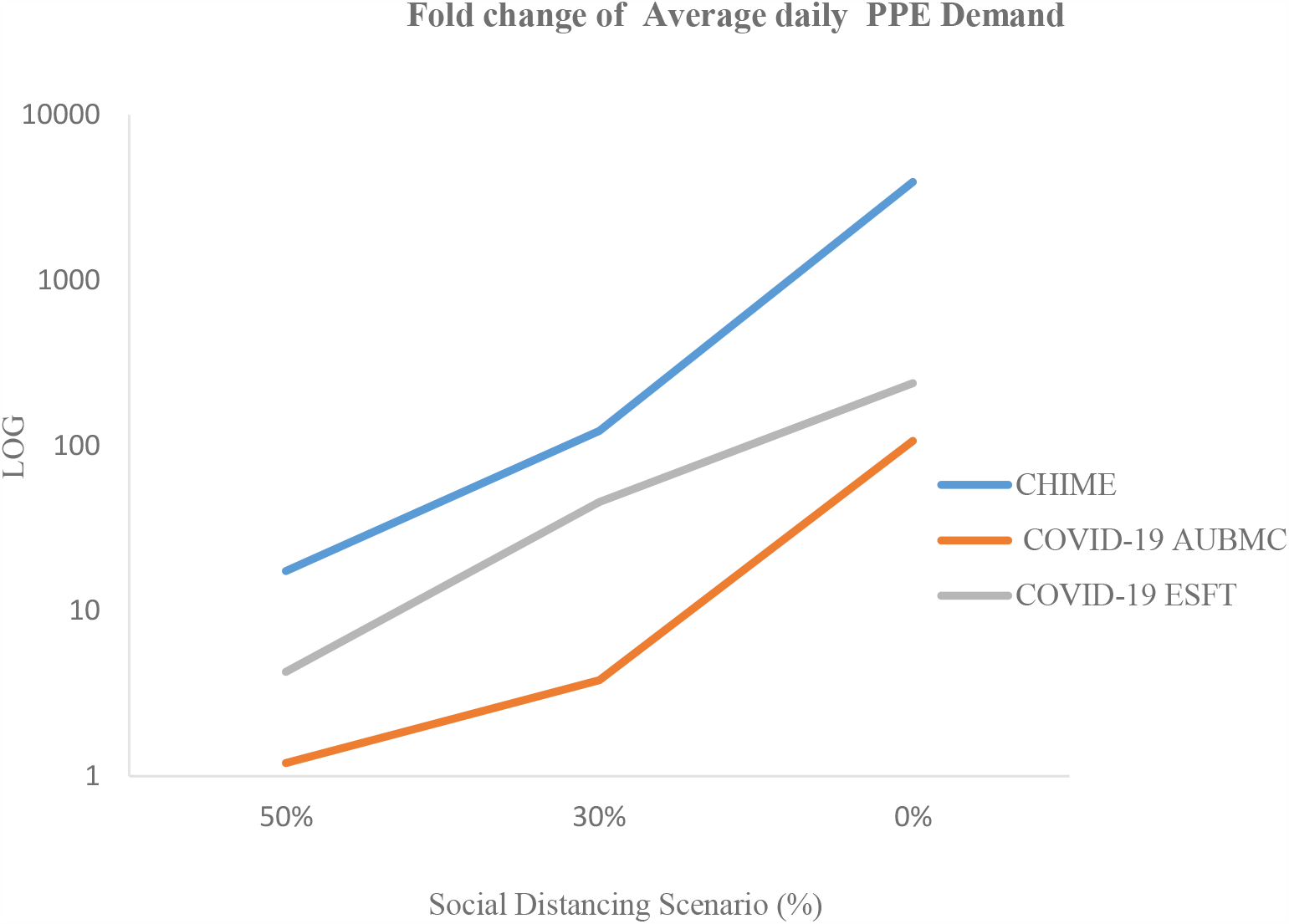
The fold change in the forecasted average daily demand for all PPE types by the three surge calculators at the 50%, 30%, and 0% social distancing scenarios compared to the Wuhan Style scenario (92%). All surge calculators used herein show a sharp increase and an excess in the average daily PPE demand upon relaxing social distancing policy measures during the COVID-19 pandemic in Lebanon up to 200 days.

## Discussion

As the first wave of the COVID-19 pandemic sweeps across the Middle East and the world, with evidence of a second wave emerging in some countries, the strain placed on the health system services including hospital beds, ICU beds, and ventilators continues to escalate. To support the preparedness for a pandemic outbreak, the capability to forecast the potential spread of a disease is an utmost need for applying public health interventions and effectively allocating resources^29^. Particularly, this is critical for low- and middle-income countries, as they often unevenly bear the infectious diseases burden and are hindered by limitations in resources available to tackle them ^29^, often relying heavily on an increasingly restricted foreign supply-chain. Despite the efficacy of forecasting in the better planning ahead and reducing the impact of the infectious disease outbreaks including healthcare capacity, deaths, and the economic burden experienced, comparing forecasts at the national level remains challenging ^17,29,30^. The latter can potentially limit the development and use of forecasting ^17^.

In this study, we highlight this challenge by comparing the projected incremental demands of medical resources and supplies by three surge calculators (CHIME, WHO COVID-19 ESFT, and COVID-19 AUBMC) for a single 200-day COVID-19 wave taking Lebanon as our case study; assuming different social distancing policy scenarios. Our results indicate that there is a discrepancy in the forecasted data between the three surge calculators despite the use of identical input data. Yet, the three surge calculators used consistently show that relaxing social distancing policy and mitigation measures can dramatically overwhelm hospital capacity and lead to a dramatic surge in the daily PPE demands during the COVID-19 pandemic. These results are congruent with recommendations for implementing initiatives to “flatten the curve” and avoid healthcare systems from being overwhelmed through the exponential growth of the disease^31^. Since prompting social distancing policy measures have helped in reducing the pandemic’s spread as shown by COVID-19 mathematical models ^32,33^, our comparison was more based on testing the efficacy of this approach on the healthcare infrastructure capacity and PPE demand using the three aforementioned surge calculators.

When assuming the strict social distancing measures (Wuhan-style), the three surge calculators projected an unpronounced daily demand for hospital beds, ventilators, and PPE during the pandemic. The variability in estimations between the calculators revealed the least margins of error given this scenario. When relaxing the social distancing measures to 50%, the calculators forecasted that the available hospital beds, ICU beds, and ventilators are enough to accommodate the projected COVID-19 cases, yet with a very noticeable increase in the average daily PPE demand. By decreasing the social distancing policy to 30%, the calculators estimate that there is a substantial surge in the demand for PPE and hospital utilization. At this stage, the difference becomes more apparent between the forecasted data. However, when the simulated social distancing measures are completely removed, all calculators show to some extent that the COVID-19 pandemic will put unprecedented strain on inpatient beds, ICU beds, ventilators, and there will be a drastic increase in the average daily PPE demands. This underscores the urgency of implementing social distancing to help in limiting COVID-19 community transmission by reducing the circle of social contacts and the contact rate ^34^. Our results are similar to those of the IHME team and Murray who forecasted that the healthcare system’s capacity will be stressed in the USA and Europe during the COVID-19 pandemic and proposed some measures to increase the supply of key products and services^8,9^. Interestingly, our data indicate that the output predictions of all used surge calculators vary very widely upon relaxing the social distancing policy measures leading to a rise in the margins of agreement.

The discrepancy in the obtained results could be related to several reasons. For PPE, some of the items are subdivided into different categories such as the types of gloves, gowns, and masks used by patients and staff as in the WHO COVID-19 ESFT calculator. Also, the ESFT calculator gives a more detailed quantification for PPE, for example, the estimation for gloves refers to the sum of those used for surgery, examination, and heavy duties. In the CHIME and COVID-19 AUBMC calculators, some of these categories are missing, which indicates that the PPE types are not being included in the calculated demand. In the COVID-19 AUBMC calculator, the N95 masks are considered to be used only in intubations. However, the ESFT calculator does not even give estimates for N95 masks. Moreover, the CHIME and ESFT calculators include PPE needs for testing staff and HCW, which adds to the demand in its estimations. Considering the need for PPE in healthcare departments related to non-COVID-19 patients and used for additional precaution could also increase the calculated demand. Varying results across the three calculators are also evident in the inpatient beds, ICU beds, and ventilators needed during the peak demand period. This can be related to the way each calculator assesses the severity of the cases. According to the COVID-19 AUBMC calculator, each ICU patient is assumed to require ventilation. However, the WHO ESFT calculator assumes that only severe and critical COVID-19 cases are admitted to the hospital and assumes that only the critical cases in ICU require ventilation, while the rest need oxygen tubes. As for the CHIME, it classifies the hospital admitted COVID-19 into three categories including hospitalized, ICU, and ventilated. Since there is no absolute truth that we can compare to assess the accuracy, we only resort to the relative comparison. Nevertheless, this wide variability in estimations can add more uncertainty to the forecasted hospital utilization and PPE demand and highlights the importance of maintaining social distancing in the absence of pharmaceutical intervention for COVID-19. To be able to evaluate the surge calculators objectively, the results produced should be compared with their real known values. However, parameters used in models for forecasting the dissemination of infectious diseases are prone to uncertainties and limitations ^29^. Besides, enhancing the current model forecasting abilities is directly proportional to the accuracy of the data provided ^29^. In our case, we input the cumulative projected COVID-19 cases as obtained from the modified CHIME model application into all calculators to maintain consistency. Yet, in addition to using already built models, we still lack accuracy as we do not have tangible data in Lebanon on PPE consumption and the capacity of healthcare system infrastructure. Also, we are not very sure how much our input data are up-to-date and reliable especially that the healthcare capacity is subjected to change throughout an outbreak ^29^.

The results of this study were based on showing the differences in estimations done by three surge calculators for COVID-19 and a modified version of the CHIME model to measure needs for different health system resources based on the total of predicted simultaneously active cases. These calculations were carried out based on three important assets receiving significant attention worldwide: inpatient beds, ICU beds, ventilators, and PPE ^35,36^. Other items and human resources required in the diagnostic and treatment chain can be forecasted including staff and HCW in the frontline of COVID-19 response, and therapeutics for supportive treatment. We believe that one of the major limitations for forecasting COVID-19 is based on the limited evidence since neither the magnitude nor duration of the COVID-19 wave is known with certainty. Another limitation is that we did not factor in PPE re-use measures such as sterilization of used N95 masks. Therefore, the adequate management of medical resources (including PPE, beds, ventilators, and health care providers) is highly recommended at this stage as some countries have started to experience a resurgence of COVID-19 cases as the pandemic continues to accelerate. Experience from several countries including China, South Korea, and Singapore in addition to mathematical modeling has revealed that the pandemic can be contained even in the exponential growth phase using a combination of interventions^34,37^. The latter mainly includes social and physical distancing, public awareness, and wearing masks ^34^.

In conclusion, the surge calculators used here, regardless of the variability in outputs, can be powerful tools for measuring the impact of social distancing policy through highlighting the dangers of scaling down non-pharmaceutical public and social health measures in the absence of any vaccine or therapy against COVID-19. Characterizing forecasting uncertainty can be improved by some promising avenues including methodological advances in model comparison and averaging ^29^. Therefore, the use of more than one model is recommended to generate more accurate and better predictions of the pandemic’s evolution ^38^. In other words, policymakers can use these calculators interconnected with each other based on the available data for each country to understand the pandemic from all its angles to be able to generate policymaking frameworks. This urges the need for a clear methodology that allows policymakers to decide which model is more applicable or adaptable for their context ^17^, and underscores the necessity of enabling calculators to be adopted to local policies and behaviors beyond social distancing. Although gaps in the present data streams provide a challenge for the current epidemic forecasting, recent breakthroughs in this field afford the possibility for refining future predictive models ^29^. Since the most essential piece of the puzzle in forecasting is data or the quality of data source, we suggest that should the data be more accurate, one can provide pandemic forecasting with fewer constraints.

## Data Availability

Requests for access to the data that support this study should be made to the corresponding Authors Dr. Hussain Ismaeel and Dr. Imad Elhajj.

## Acknowledgement

None

## Financial Support

None

## Conflict of interest

No competing interest declared.

## Author’s Contribution

AK and NM have contributed to the design and implementation of the research, to the analysis of the results, and the writing of the manuscript. EH and GE contributed to data collection. IE and HI conceived the study and were in charge of overall direction and planning. All authors discussed the results and contributed to the final manuscript.

## Data Availability Statement

Requests for access to the data that support this study should be made to the corresponding Authors: Dr. Hussain Isma’eel and Dr. Imad Elhajj.

## Notes

### Competing Interest Statement

The authors have declared no competing interest.

### Funding Statement

No funding available

### Author Declarations

No IRB needed since there are no human subjects used

